# Artificial Intelligence-Based Modeling to Aid in the Identification of Genetic Conditions in Infants with Congenital Heart Disease

**DOI:** 10.64898/2026.10.12.26365328

**Authors:** Michael Caiola, Tyler Thompson, Sara B. Stephens, Bita Salamat, Krista Harris, Claudia Lara, Zeynep Coban Akdemir, Lilei Zhang, Matthew Roth, Aleksandar Milosavljevic, Andrew C. Weitz, Shaine A. Morris

## Abstract

**Background:** Genetic conditions are common in congenital heart disease (CHD) and affect care, but are often missed or diagnosed late. We asked whether routine clinical data can shortlist the most likely genetic conditions.

**Methods:** Retrospective single-center cohort of children with CHD and an echocardiogram who had either a confirmed genetic diagnosis or negative cytogenetic testing and CHD gene sequencing. Echocardiogram images and serial medical record data were added as model inputs in the order they become clinically available. Class-balanced logistic regression (LR) and a multimodal deep learning fusion network were fitted to the same inputs at each step. The primary metric was balanced top-3 accuracy (correct diagnosis among the top three, averaged across conditions), with bootstrap intervals.

**Results:** The cohort included 1,720 children (988 with common CHD-associated genetic conditions, 231 rare diagnoses, 501 negative testing). Using only inputs generated at the echocardiogram visit, LR reached a balanced top-3 accuracy 0.758 (chance 0.429). Serial records, radiology notes and ICD-10 codes, raised this to 0.809. Multimodal fusion performed similarly at each stage. With a genetic condition withheld from training, the fusion embedding placed affected children nearest to others sharing a gene group in 0.37 of cases versus 0.10 without the embedding (+0.268 [95% CI +0.242, +0.295]).

**Conclusions:** Routine clinical data available at first echocardiogram can help identify likely genetic diagnoses, and later coded inputs ceiling. Multimodal fusion added no accuracy over LR, but places children sharing a genetic cause near one another, making nearest-neighbor retrieval informative for undiagnosed children.

## INTRODUCTION

Congenital heart disease (CHD) is frequently the first presenting sign of an underlying genetic condition, with a genetic etiology reported in 20-30% of CHD cases when broad testing is employed^1,2^. Recognizing a genetic condition can dramatically alter prognosis, surveillance, acute perioperative and chronic management, and family counseling^1^. However, genetic testing algorithms among newborns with CHD are highly variable by center. A recent multi-institutional survey of pediatric cardiac intensive care units demonstrated that only 58% of respondents obtained routine genetic testing for all affected neonates with CHD and only 28% obtained a routine formal genetics consultation^3^. This heterogeneity was most pronounced for isolated CHD, where testing was offered routinely by just 46% of institutions and first-line test selection differed widely across respondents^3^. Some recent studies have studied universal broad testing (e.g. whole genome sequencing) in CHD, which demonstrates a high yield with relatively quick return of results^4–6^. However, this strategy is not possible in most clinical settings at this time, often limited by cost of testing and by genetic professional staffing, as broad testing requires detailed pre-test counseling and informed consent.

Artificial-intelligence (AI) phenotyping has been explored as a tool to improve genetic diagnosis and target appropriate genetic testing. For example, facial-image analysis identifies 22q11.2 deletion, Williams, and Noonan syndromes, as well as vascular Ehlers–Danlos syndrome from a handful of photographs with high accuracy^7–11^. However, facial AI tools are typically developed in and perform best beyond the newborn period, because the characteristic facial gestalt of many syndromes is often subtle at birth and becomes more recognizable with postnatal development, potentially limiting their utility in newborns^12–14^. In contrast, abnormal cardiac morphology is present and stable from the prenatal period (when prenatal imaging is performed), and universally at the time of cardiac diagnosis with echocardiographic images.

We hypothesized that echocardiographic images would provide rich information to aid the identification of known genetic conditions which then would be further strengthened by multi-modal clinical input. Phenotype matching is an established strategy for genetic diagnosis. For example, Matchmaker Exchange is a federated network connecting databases of genomic and phenotypic data that facilitates the identification of patients with overlapping phenotypes and variants in shared candidate genes, supporting the discovery of common genetic etiologies^15^. Quantitative, imaging-informed phenotyping has demonstrated value for matching patients with shared facial imaging features within defined genotype groups in other systems^8,9^ and prioritizing causal genes in inherited retinal disease^16^. An important next step is to determine whether integrating detailed, echocardiographic and complementary clinical data aid in etiologic patient matching. In the present study, we create and compare two multi-modal models in children with CHD to predict genetic diagnosis using echocardiographic images, echocardiogram reports, Fyler codes, and other records extracted from the electronic health record including demographics, International Classification of Disease (ICD) 10^th^ edition codes, and growth parameters.

## METHODS

### Study Population

This retrospective single tertiary center cohort included children evaluated at Texas Children’s Hospital from January 1, 2010 to March 31, 2026 with a clinical diagnosis of CHD, ≥1 postnatal echocardiogram prior to invasive cardiac interventions with images available for review, and either a 1) confirmed genetic diagnosis or 2) nondiagnostic comprehensive genetic testing. Comprehensive genetic testing was defined as one of the following: 1) chromosomal microarray and whole exome sequencing (proband or familial), 2) chromosomal microarray and CHD or heterotaxy gene sequencing panel, or 3) whole genome sequencing. In cases where a genetic diagnosis was made that was not thought to be the etiology of the CHD, these were still considered as negative testing in regard to CHD. Individuals with isolated cardiomyopathy or arrhythmia disorders without structural CHD were excluded. As well, individuals with only a patent foramen ovale or patent ductus arteriosus in the newborn period were excluded. Subjects were included under a retrospective waiver of consent as approved by the Institutional Review Board at Baylor College of Medicine (H-57914).

### Data Collected

Data collected included: Echocardiographic DICOM images (first comprehensive study, 1 study per individual), the corresponding echocardiogram report (including tabular data and free text) and diagnostic codes assigned by signing cardiologist (Fyler codes), primary CHD diagnosis, demographics, radiology reports, and ICD-10 diagnostic codes corresponding to structural birth defects (excluding genetic conditions). Primary CHD diagnosis was ascertained by two pediatric cardiologists (S.A.M., B.S.). Demographics evaluated included sex and age at echocardiogram. Primary height (in centimeters) and weight (in kilograms) at time of echocardiogram, in addition to serial height and weight measurements indexed by age, were also collected. Echocardiographic and other radiologic reports (e.g., cardiac magnetic resonance imaging (cMRI), computed tomography (CT), and non-cardiac ultrasound) were included as free text. Inputs into the model were added in an order to mimic clinical availability (see below). All data were deidentified and coded, including DICOM data, to remove all private health information including dates of service before entering into modeling.

### Target/Outcome

The primary target of was the genetic condition. Given that some conditions were very common, and some only occurred once in the cohort, individual were stratified according to genetic diagnosis with groupings as follows: 1) *common-7*: trisomy 21, 22q11.2 deletion syndrome (i.e. DiGeorge syndrome), Williams syndrome, trisomy 18, Turner syndrome, trisomy 13, and Kabuki syndrome; 2) *rare diagnosis*: a confirmed molecular diagnosis outside the common-7; and 3) *undiagnosed*: testing without an identified genetic etiology. To maintain adequate sample sizes, *ELN*-mediated elastin arteriopathy was included in the Williams class. *CHD7*-associated CHARGE syndrome was retained in the rare diagnostic pool.

### Model Inputs of Data Collected

1. Echocardiographic images: Per-clip embeddings from a video encoder pre-trained on adult echocardiography (EchoPrime)^17^. Preprocessing consisted of setting the pixels outside the ultrasound sector to zero, averaging over a study’s clips and reducing to 64 dimensions by truncated singular value decomposition (SVD).
2. Demographics: Sex, age at the first echocardiogram, and height, weight and body-mass-index (BMI) z-scores computed from measurements taken at or before that echocardiogram (median over that window); children with no measurement at or before the echocardiogram were median-imputed inside each training fold, and their later measurements were not used, scored against the WHO Child Growth Standards^18^ below 24 months of age and the CDC 2000 growth reference^19^ at or after 24 months, following CDC guidance for children in the United States.
3. Echocardiographic measurement z-scores (**Supplemental Table 1**): Z-scores were extracted from the echocardiographic report and were calculated using the Boston Children’s nomograms^20–22^.
4. Echocardiogram free-text report narrative, with syndrome and gene names masked (the mask list accompanies the code), as term frequency-inverse document frequency (TF-IDF) reduced to 64 dimensions by truncated SVD.
5. Echocardiogram diagnostic codes (also known as Fyler codes) reported as structured diagnostic codes assigned by the pediatric cardiologist at the time of each echocardiogram report and pooled across the child’s initial echocardiograms the same block the fusion model receives. The Fyler system is a granular, anatomy-based classification developed at Boston Children’s Hospital, using a vocabulary of >3,000 phenotypic descriptors to code congenital cardiac malformations for computerized storage, retrieval, research, and outcomes analysis^23^.
6. Radiology reports: free-text clinical impressions from cross-sectional imaging (e.g., cMRI and CT) accumulated over the record, available for 35% of the common-7 cohort, processed as in input 4 (masked, TF-IDF, 64-dimension truncated SVD) with an added indicator of whether any note existed and its log length, so that children without notes were represented rather than zero-filled.
7. Hierarchical ICD-10 birth-defect codes (Q00–Q79; Q80–Q99 were excluded due to the potential of revealing the underlying genetic condition) which were available only as an undated cumulative diagnostic record and therefore may reflect diagnostic workup performed after the echocardiogram. The radiology reports and the ICD-10 codes are the two inputs that accrue after the echocardiogram visit; all others are generated at the visit.

### Model Building and Evaluation

We first 1) evaluated a logistic regression (LR) echocardiogram-based model with additional multimodal inputs in the order inputs become clinically available, treating the lifetime diagnostic record as an upper bound, 2) compared its performance with that of a multimodal fusion model using the same cumulative inputs after each new data source was added, and 3) tested whether the fusion embedding recognizes a genetic condition absent from training as unfamiliar and places children with presumed shared genetic etiologies near one another, a question a closed-set classifier (such as LR) cannot answer (**Figure 1**).

**Figure 1.**
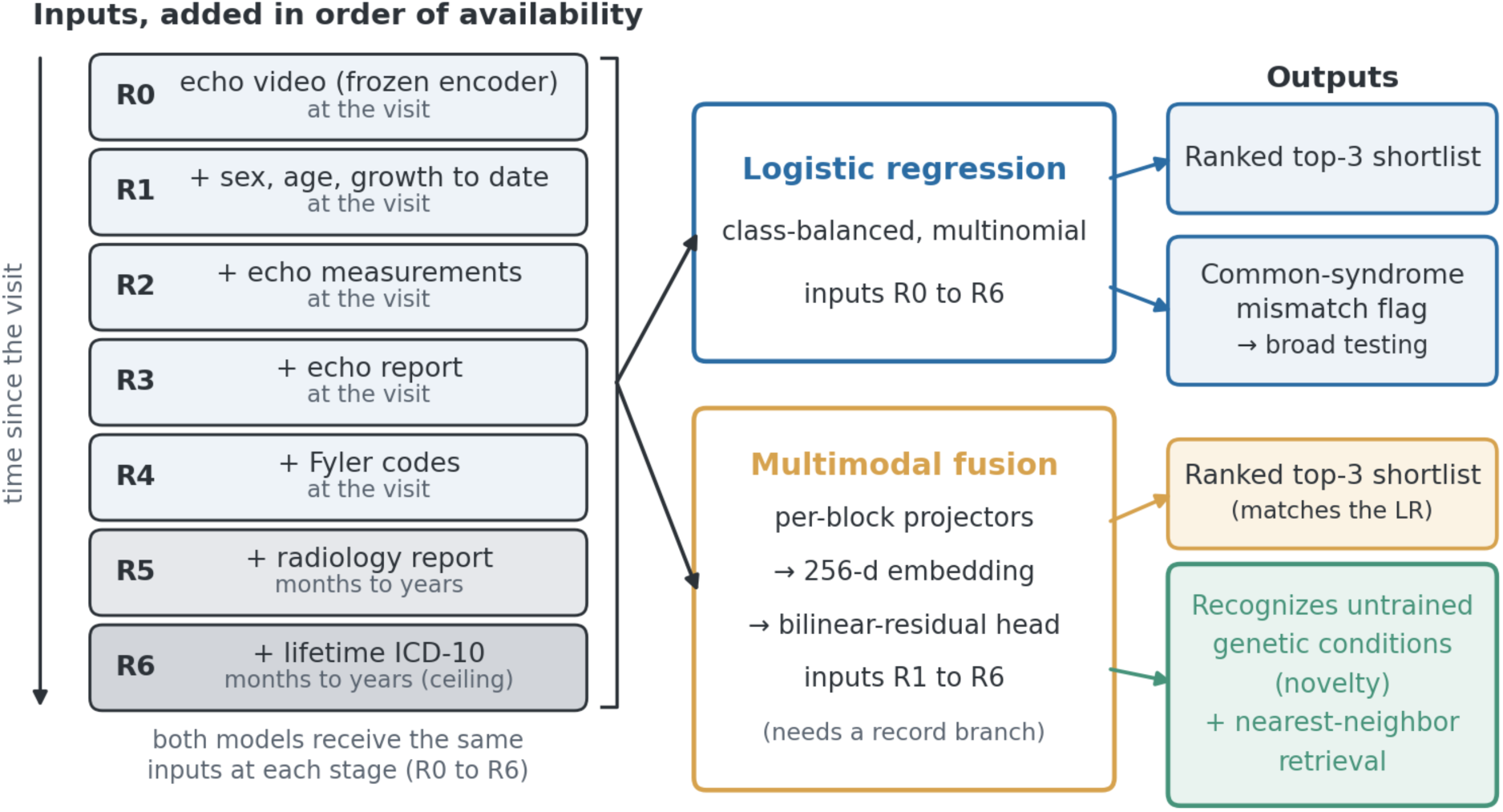
Study Overview. Study design. Inputs enter in the order they become available: the echocardiographic DICOM images (R0); sex, age, and height, weight, and BMI z-scores to the study date (R1); the echocardiographic measurements (R2); the narrative echocardiogram report (R3); and the Fyler codes from the initial echocardiograms (R4), all generated at the visit, then the radiology reports (R5) and the lifetime ICD-10 codes (R6) that accrue afterwards, shaded. Right: at each stage both models receive the same inputs. The LR returns a ranked shortlist and the common-syndrome mismatch flag; the fusion’s 256-d embedding is evaluated on genetic conditions withheld from training and on nearest-neighbor retrieval.

### Logistic Regression Model

The LR model was multinomial with L2 regularization at the default strength (scikit-learn 1.8, C = 1), class-balanced sample weights, one-hot encoding of categorical inputs, median imputation and standardization, all fitted inside each training fold; no hyperparameter was tuned. A single penalty strength cannot work with both a 69-feature and a 596-feature model; therefore, adding inputs is confounded with changing how strongly the model is regularized. As a sensitivity analysis, the penalty strength was instead selected inside each training fold by 3-fold cross-validation over six values from 0.01 to 3.

### Multimodal Fusion Model

The multimodal fusion projected each input block to 256 dimensions (linear layer, batch normalization, ReLU), combined them with a bilinear-residual layer into a shared 256-d embedding, and classified with class-weighted cross-entropy (AdamW, learning rate 10⁻³, weight decay 10⁻⁴, batch 64, 80 epochs, no early stopping; Figure 1); its report vocabulary was fitted inside each training fold. Both models were fitted to identical inputs at each stage of data availability. From the radiology report stage, the masked notes were appended to the echo report text branch, and the presence indicator and log length joined the echo record block.

### Model Evaluation

Evaluation used stratified 5-fold cross-validation repeated over three seeds. The primary metric was balanced top-3 accuracy. “Top-3” reflects that the model returns a ranked shortlist rather than a single forced diagnosis, and a diagnosis counts as correct if it appears among the three. “Balanced” reflects that each syndrome is weighted equally, so that conditions comprising the majority of the cohort cannot dominate the average. For each of the common-7 syndromes, the proportion of children with that syndrome whose diagnosis the model ranked in the top 3 was computed. The metric was the average of those seven proportions (chance 3/7 = 0.429), the top-k form of balanced accuracy^24^.Every reported accuracy carried a 95% subject-level bootstrap confidence interval (CI) (20,000 resamples, stratified within class, seeds averaged inside each replicate). Every comparison between arms was paired on the same subjects, and adjacent inputs were compared as paired differences. The three truncated-SVD reductions used by the LR (64 components each, of the pooled echocardiogram embedding, of the report TF-IDF and of the radiology report TF-IDF) were fitted on all 988 common-7 subjects without labels; this is the one place information crosses folds, and it is label-free. Splits were grouped by subject but not by echocardiography scanner. The fusion requires a record branch and cannot run on the images alone, thus, its evaluation begins when demographic and growth data are added.

### Syndromes Withheld from Training and the Common-Syndrome Mismatch Flag

The primary shortlist analysis included the common-7 syndromes. For a separate leave-one-group-out analysis, we expanded the class set to 11 diagnostic genetic syndrome groups by adding RASopathies (n=15), ciliary/laterality disorders (n=17), CHARGE syndrome (n=17), and Alagille syndrome (n=7); the gene-level membership of the merged groups is given in **Supplemental Table 2**. In each iteration, one group was withheld entirely, and the fusion embedding was trained on the remaining 10 diagnostic groups with 25% of those patients reserved for validation. This analysis used a gated fusion head and per-view echocardiographic features rather than the bilinear-residual head and pooled features used for the shortlist, as the gated variant is the stronger of the two on this task (unfamiliar-condition detection +0.049, 95% CI +0.021 to +0.077). Removing the echocardiographic branch entirely changes the same-cause neighbor and recall measures by less than 0.01, thus, the difference in echocardiographic representation does not affect the comparison. We then calculated three embedding-based measures and compared them with the same measures derived from the identical inputs concatenated without the learned embedding, so that the comparison isolates the representation rather than the information available to it. Because a flat concatenation is weakened by the nine per-view echocardiographic blocks, which outnumber the five record blocks, we also report each comparison against the strongest raw baseline, which omits the echocardiogram. The first, unfamiliar-condition detection, decided whether the embedding separated patients with the withheld syndrome from children with validated genetic syndromes it was trained on; it is scored as the area under the receiver operating characteristic curve (AUROC) of each child’s highest similarity to any trained-class centroid^25^. The second, same-cause neighbors, was computed among children with rare genetic conditions that no model was trained on (n=121 per iteration: every rare child in a confirmed gene group of two or more members): the fraction whose nearest neighbor in the embedding has a genetic condition in the same gene group, where rare conditions are grouped by causal gene or pathway (e.g., the RAS-pathway genes). Chance for this measure, the probability that a random other rare child shares the query’s gene group, is 0.034. The third, recall on the known classes, is nearest-centroid recall@1 on the validation patients and is a floor. Each contrast was averaged over seeds within group and reported with a t-interval across the 11 groups, as the 33 iterations are 11 groups × 3 seeds rather than 33 independent draws.

The common-syndrome mismatch flag indicates that a child does not fit any of the common-7 syndromes. It was a supervised detector: a LR separating the common-7 group from the undiagnosed group, with rare diagnosis patients held out, trained by 5-fold cross-validation over three seeds and scored out of fold. We report the AUROC against the undiagnosed group and against the rare group, with subject-level bootstrap intervals (20,000 resamples). The detector was fitted on two input sets: the at-visit inputs available through completion of the echocardiogram report (sex, age, height, weight and BMI z-scores, the echocardiographic measurement z-scores including blood pressure, echo report free text, and DICOM images) and the coded inputs (the same sex, age, height, weight and BMI z-scores, and the lifetime ICD-10 codes). The Fyler codes, although generated at the visit, were omitted from the at-visit detector because they did not improve it: adding them lowered the AUROC against the undiagnosed group from 0.896 to 0.846 at the fixed penalty and to 0.894 when the penalty was selected inside each fold, against 0.907 without them. Height, weight and BMI z-scores are the at-or-before-echo values defined under Model Inputs throughout, including the record block of the leave-one-group-out analysis. One label-free distance was scored the same way: distance to the nearest trained-class centroid in the fusion embedding. Analyses were performed using custom Python scripts.

To visualize the embedding, the 256-dimensional fusion embedding with every input was projected to two dimensions by UMAP^26^ (cosine metric, 20 neighbors), once from the model trained on all seven syndromes and once from a single cross-validation fold, so that in the second layout every plotted child was unseen by the model that embedded it.

## RESULTS

### Cohort Composition

In total, 1,720 patients were included in the final model, including 1,219 with a genetic diagnosis and 501 with non-diagnostic testing (70.9% and 29.1% respectively; **Table 1**). Of those with a genetic diagnosis, 988 patients were included in the common-7 diagnostic group, including 580 with trisomy 21, 152 with 22q11.2 deletion syndrome, 91 with Williams syndrome and *ELN* variants, 67 with trisomy 18, 45 with Turner syndrome, 34 with trisomy 13, and 19 with Kabuki syndrome (**Supplemental Table 3**). The remaining 231 patients were classified as having a rare diagnosis.

**Table 1.**
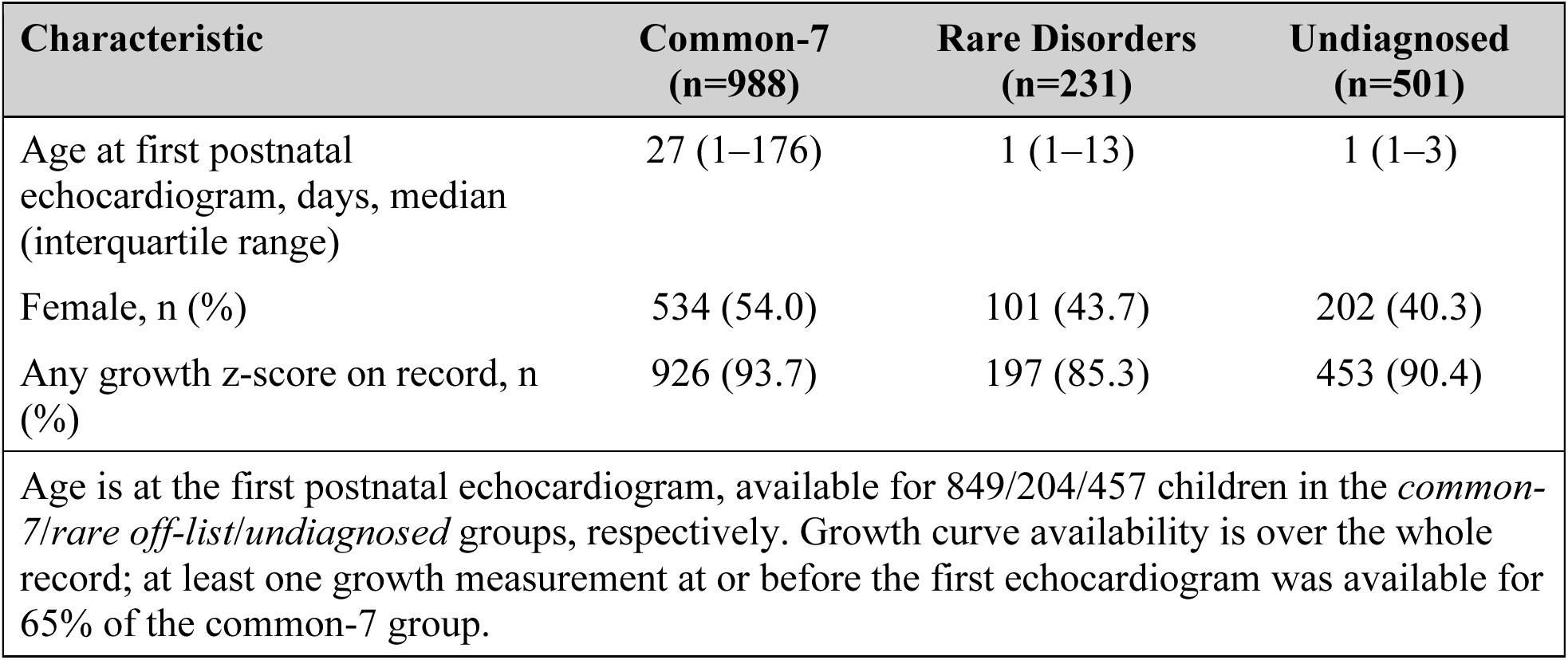
Cohort Characteristics by Diagnostic Group.

### First Echocardiographic Data

The echocardiogram images alone ranked the common-7 syndromes at a balanced top-3 accuracy of 0.611 (95% CI 0.576 to 0.649; **Table 2**); namely, on average across the 7 classes, the model identified the correct syndrome in its top 3 predictions in 61.1% of patients. Adding the other inputs generated at the echocardiogram visit changed it in turn: sex, age and height, weight, and BMI z-scores to 0.642 (95% CI 0.606 to 0.680; +0.031; 95% CI +0.004 to +0.058), the echocardiographic measurements to 0.655 (95% CI 0.620 to 0.691; +0.013; 95% CI -0.017 to +0.043; **Figure 2B**), the echocardiogram report to 0.761 (95% CI 0.723 to 0.799), the largest single increment (+0.106 (95% CI +0.072 to +0.140)), and the Fyler codes to 0.758 (95% CI 0.721 to 0.796; -0.002; 95% CI -0.011 to +0.006). The two inputs that accrue after the visit raised accuracy to 0.809 (95% CI 0.771 to 0.846), a gain of +0.051 (95% CI +0.018 to +0.084) over the at-visit model: the radiology reports contributed +0.020 (95% CI - 0.011 to +0.053) and the lifetime ICD-10 codes +0.030 (95% CI +0.009 to +0.053).

**Figure 2.**
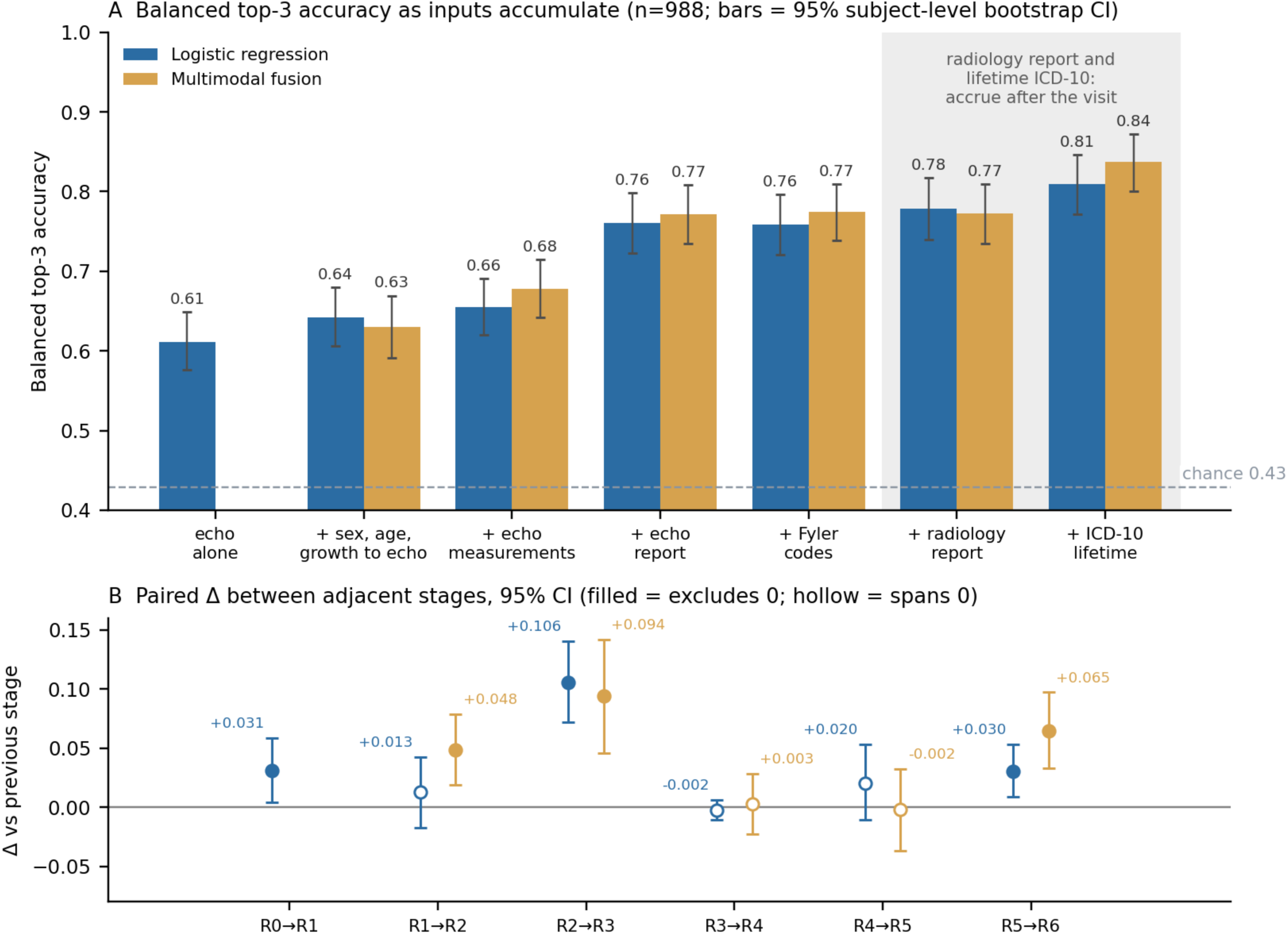
Model Performance with Cumulative Input. Echo: Echocardiogram. (A) Balanced top-3 accuracy of the LR (blue) and the multimodal fusion (orange) as inputs accumulate, one block at a time; bars are 95% subject-level bootstrap CIs (n=988); shaded input sets (radiology reports, ICD-10) accrue after the visit; dashed line, chance (0.429). (B) Paired difference between adjacent input sets with 95% CI; filled markers exclude zero.

**Table 2.** Balanced Top-3 Accuracy with Cumulative Input, Stratified by Model Type.

| **Inputs available** | **LR** | **Fusion** | **Fusion – LR** |
| --- | --- | --- | --- |
| Echo alone | 0.611 (0.576 - 0.649) | — | — |
| + Sex, age, height, weight, and BMI (at echo) | 0.642 (0.606 - 0.680) | 0.630 (0.591 - 0.669) | -0.013 (-0.053, +0.029) |
| + Echo measurements | 0.655 (0.620 - 0.691) | 0.678 (0.642 - 0.715) | +0.023 (-0.016, +0.062) |
| + Echo report | 0.761 (0.723 - 0.799) | 0.772 (0.735 - 0.809) | +0.011 (-0.034, +0.056) |
| + Fyler codes | 0.758 (0.721 - 0.796) | 0.775 (0.738 - 0.810) | +0.016 (-0.024, +0.056) |
| + Radiology reports | 0.779 (0.740 - 0.818) | 0.773 (0.735 - 0.810) | -0.006 (-0.050, +0.040) |
| + ICD-10 lifetime codes | 0.809 (0.771 - 0.846) | 0.837 (0.800 - 0.872) | +0.028 (-0.016, +0.073) |
| Echo: Echocardiogram; ICD: International Classification of Disease; LR: Logistic regression; Balanced top-3 accuracy for chance is 0.429. Scores reported are balanced top-3 accuracy with 95% bootstrap confidence intervals. |  |  |  |

Decomposed, an indicator of whether any radiology report existed, together with its length, contributed +0.036 (95% CI +0.010 to +0.064) on its own, while the note text contributed +0.015 (95% CI -0.015 to +0.047) and, combined with the indicator, less than the indicator alone. Notes were present for 74% of children with Kabuki syndrome and 59% with 22q11.2 deletion syndrome but 28% with trisomy 21 and 21% with Williams syndrome, so the presence of cross-sectional imaging is itself a marker of the workup a child received. Once the ICD-10 codes were present, the notes did not add any additional accuracy (-0.002; 95% CI -0.026 to +0.022).

Adding blocks did not always improve LR performance, as the model with exclusively coded inputs (sex, age, height, weight, and BMI z-scores, and lifetime ICD-10 codes, without the imaging-derived inputs) scored a balanced top-3 accuracy of 0.891 (95% CI 0.855 to 0.924), exceeding the model with all inputs by +0.082 (95% CI +0.039 to +0.125) and also the model with all inputs except the echocardiogram images (0.816 (95% CI 0.781 to 0.850)). This was not an artifact of model configuration. When the penalty strength was chosen inside each training fold instead of fixed in advance, the at-visit stages rose by up to 0.03, the full model changed by +0.01 and the coded-only model by less than 0.01, and the coded-only model still exceeded the full model (0.889 vs 0.816).

### Multimodal Fusion Compared to Logistic Regression

The multimodal fusion model performed similarly to the LR at each input stage (**Figure 2**, **Table 2**). Paired differences between the models (fusion-LR) ranged from -0.013 to +0.028 across the six shared stages, and none excluded zero (**Table 2**). The fusion’s stage-to-stage gains resembled the LR’s except at the measurements step, where the fusion gained +0.048 (95% CI +0.019 to +0.079) and the LR did not; its echocardiogram report step was +0.094 (95% CI +0.045 to +0.141) and its lifetime ICD-10 step +0.065 (95% CI +0.033 to +0.097). At this sample size (n=988), the fusion did not improve the shortlist over the LR.

The echocardiogram report captured most of the predictive information provided by echocardiographic DICOM imaging in the LR, and the Fyler codes captured the remainder in the fusion (**Figure 3**). For the LR model, the images improved on sex, age and height, weight, and BMI z-scores by +0.053 (95% CI +0.003 to +0.103) and again once the radiology report was present (+0.041, 95% CI +0.009 to +0.075), with nothing detectable at the other stages; with all inputs, the model without images scored 0.816 (95% CI 0.781 to 0.850) against 0.809 with them (-0.007; 95% CI -0.036 to +0.023). For the fusion, the images contributed +0.048 (95% CI +0.008 to +0.089) once the echocardiographic measurements were present and +0.030 (95% CI +0.002 to +0.059) once the report was present, and nothing detectable after that (all inputs: -0.000; 95% CI -0.025 to +0.025).

**Figure 3.**
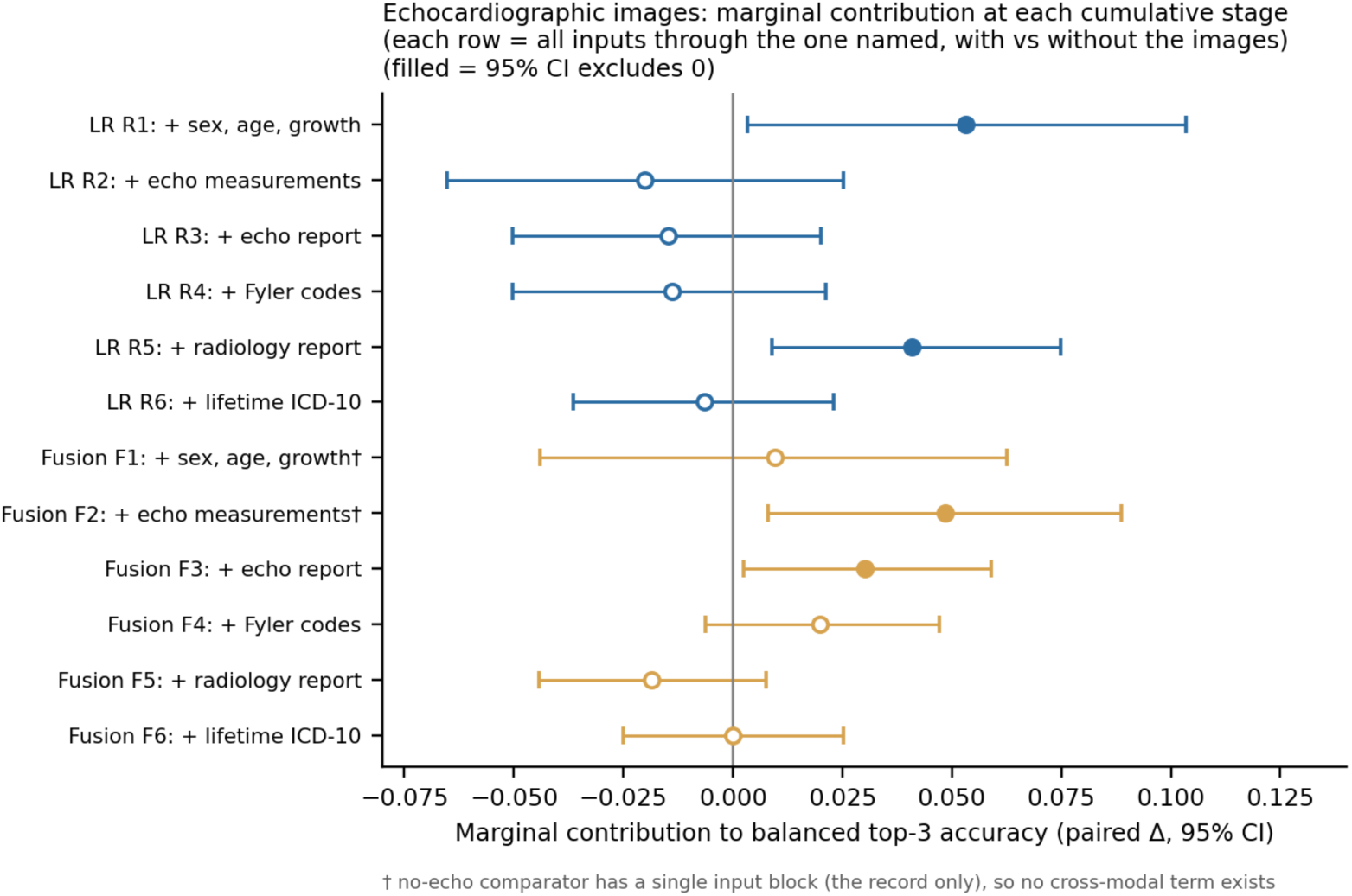
Difference in Balanced Top-3 Accuracy by Model with Cumulative Inputs. Marginal contribution of the echocardiographic images at each cumulative stage. Each row compares the full stage, every input through the one named, with and without the echocardiographic images, as a paired subject-level bootstrap Δ in balanced top-3 accuracy with 95% CI; filled markers exclude zero. Stage codes as in Figure 1. LR (blue), fusion (orange). † At F1 and F2 the no-echo comparator has a single input block and no cross-modal term.

### Recognition of Unfamiliar Genetic Etiologies and Grouping Patients with Similar Genetic Conditions

When a genetic condition was withheld from training, the embedding separated affected children from those with the trained conditions more accurately than the same inputs concatenated without the learned embedding (AUROC 0.705 vs 0.577; +0.128; 95% CI +0.066 to +0.190; **Figure 4A**). The embedding led in 11 of the 11 withheld groups. Among children with rare genetic conditions not included in model training, the nearest patient identified by the embedding shared the same gene group 36.8% of the time, compared with 9.9% for the same inputs without the embedding and 3.4% by chance (+0.268; 95% CI +0.242 to +0.295; 11 of 11 groups). The embedding also organized the trained conditions more cleanly under the same nearest-centroid rule (recall@1 0.504 vs 0.426; +0.079; 95% CI +0.049 to +0.108). Because a flat concatenation is weakened by the nine per-view echocardiographic blocks, we repeated each comparison against the strongest alternative baseline, which omits the echocardiogram: the detection advantage narrowed to +0.101 (95% CI +0.023 to +0.180) and the recall advantage to +0.052 (95% CI +0.023 to +0.080), while the gene-group neighbor advantage was little changed (+0.252; 95% CI +0.225 to +0.279). Removing the echocardiographic branch from the fusion itself changed the neighbor and recall measures by less than 0.01, so the retrieval results were not dependent on imaging.

**Figure 4.**
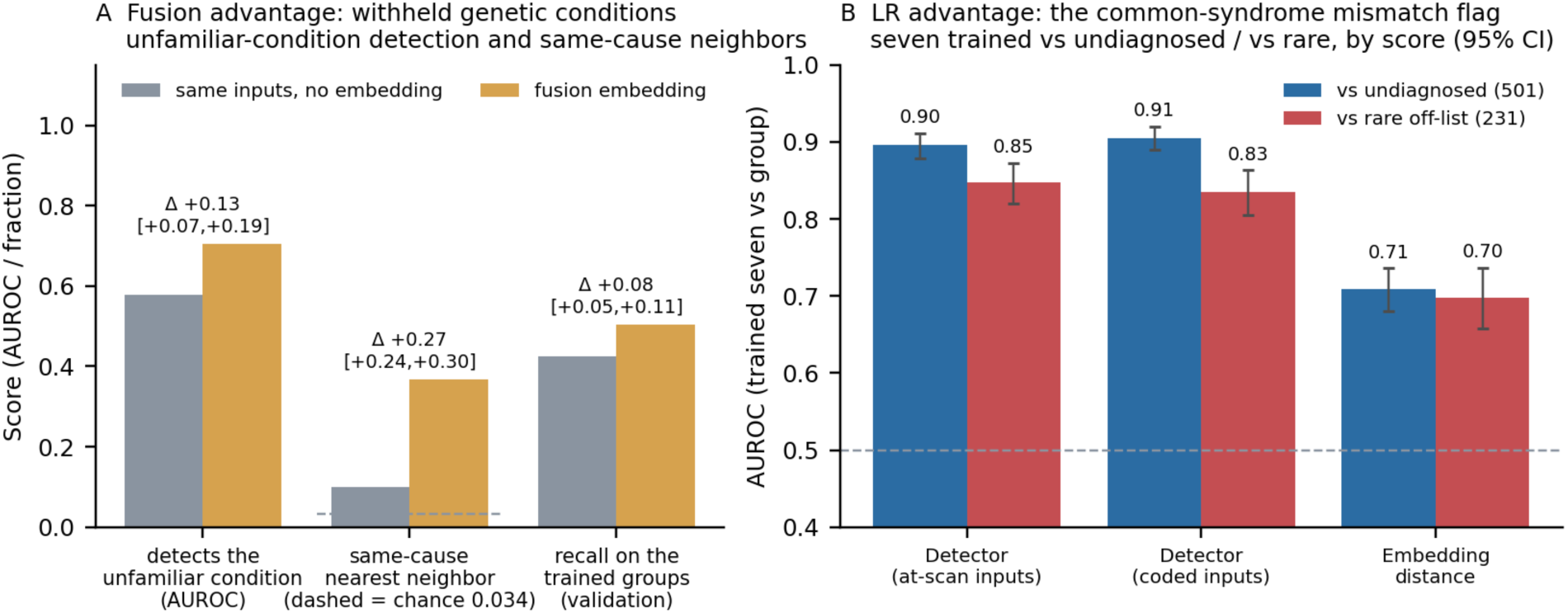
Comparison of Multimodal Fusion and Logistic Regression Model Performance. (A) Leave-one-group-out analysis: for each of 11 genetic condition groups withheld in turn, unfamiliar-condition detection (AUROC separating children with the withheld group from validation children with the trained groups, by highest centroid similarity), same-cause neighbors (children with rare conditions outside all training whose nearest neighbor shares their gene group; dashed line = chance, 0.034), and recall@1 on the trained groups for the validation children. Both arms receive the same inputs; the gray bars concatenate them without the learned embedding. Δ is shown with 95% class-level confidence intervals over the 11 groups. (B) The common-syndrome mismatch flag, with all seven common syndromes trained: AUROC separating the common-7 group from the undiagnosed group (n=501) and from rare conditions (n=231), for a supervised detector on two input sets and for distance to the nearest trained centroid in the fusion embedding. Bars are 95% subject-level bootstrap confidence intervals.

### Common-Syndrome Mismatch Flag Results

The common-syndrome mismatch flag addresses a different question from **Figure 4A**. Compared to requiring the model to name the exact genetic condition a child has, this model was only required to answer a binary question of whether the individual had one of the common-7 genetic syndromes or not. A supervised detector was able to better discern this compared to any distance in the embedding (**Figure 4B**). Using data available from the initial echo evaluation alone, the detector separated the common-7 syndromes from the undiagnosed group with an AUROC of 0.896 (95% CI 0.879 to 0.912), and from rare genetic conditions not included in training with an AUROC of 0.847 (95% CI 0.820 to 0.873). At a threshold that flagged 90% of the undiagnosed group, the model also flagged 31% (95% CI 26% to 37%) of children with a common-7 genetic syndrome and 85% (95% CI 79% to 90%) of the rare group. With the coded data instead (sex, age, height, weight, and BMI z-scores, and lifetime ICD-10 codes, without the imaging-derived inputs), the detector reached 0.905 (95% CI 0.890 to 0.920) for the common-7 and 0.835 (95% CI 0.805 to 0.863) for the rare groups. Distance to the nearest trained centroid in the fusion embedding reached 0.709 (95% CI 0.681 to 0.736) and 0.698 (95% CI 0.658 to 0.737).

### Embedding Space

**Figure 5** shows the fusion embedding from one cross-validation model. The held-out children with trisomy 21, 22q11.2 deletion syndrome and Williams syndrome fall on their syndromes’ regions while the smaller syndromes disperse, and the undiagnosed and rare children, none of whom any model was trained on, spread along bands between the syndrome regions rather than forming groups of their own, which is the picture the same-cause-neighbor and flag results describe numerically.

**Figure 5.**
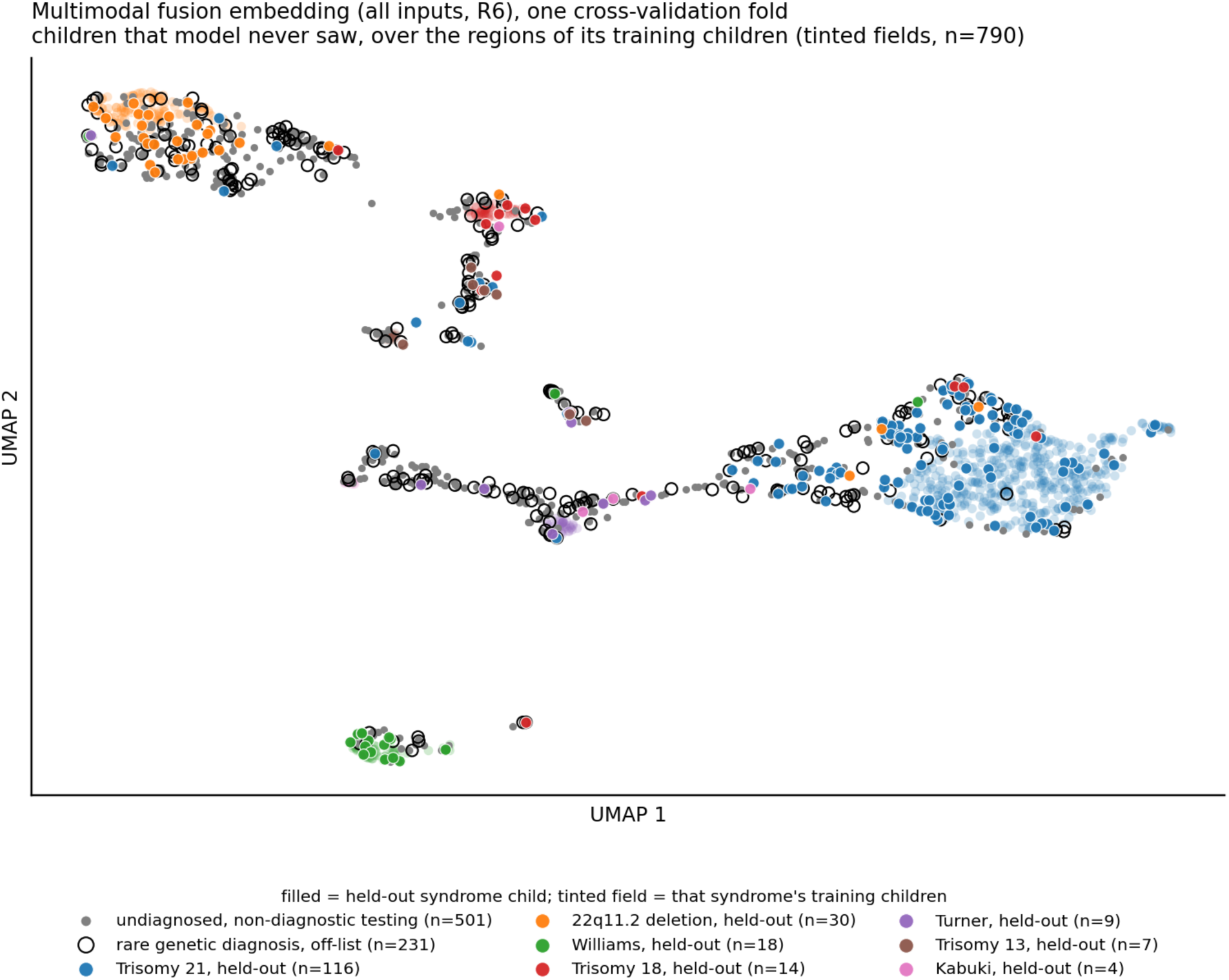
Fusion Embedding Space. The fusion embedding with every input from a single cross-validation model, projected to two dimensions by UMAP, showing the children that model never saw: the held-out 20% of each syndrome (filled, colored by syndrome), all undiagnosed children (gray) and all rare-diagnosis children (open circles). The tinted fields are that model’s training children (n=790), colored by syndrome, so each unseen child can be read against the syndrome regions the model learned. The layout was fitted once on all children embedded by that model. It is illustrative and carries no quantitative claim beyond Figure 4.

## DISCUSSION

About 20-30% of patients with CHD receive a genetic diagnosis, despite strong evidence that a substantially larger proportion have an underlying genetic etiology^27–32^. Although certain genetic syndromes are more readily diagnosed due to recognition of phenotypic features and extracardiac manifestations, these features may not be obvious after the newborn period, and an early molecular confirmation remains critical. A confirmed genetic diagnosis can directly influence clinical management, surgical decision making, surveillance protocols, and family counseling^33^. For instance, patients with Turner syndrome who have hypoplastic left heart syndrome are known to have poor outcomes following stage I single ventricle palliation^34^; consequently, some institutions may not offer palliative surgery. Other examples include the need for hematological evaluation in patients with Noonan syndrome prior to invasive interventions^35^, or the higher awareness and diligent perioperative planning due to the risk of sudden cardiac death in patients with William syndrome^36–39^. Earlier genetic diagnoses may also facilitate timely neurodevelopmental screening and intervention, especially in patients with CHD who undergo surgical intervention ^40,41–44^.

Genomics and genetic testing are rapidly advancing; however, limitations still exist. Access to genetic evaluation and testing remains highly variable across institutions. While larger tertiary referral centers often have established resources, specialized personnel, and standardized protocols to support broad and comprehensive genetic testing, smaller centers, particularly community-based programs, may lack such foundation. Even among tertiary centers, significant variability exists in both genetic evaluation and testing strategy practices^3^. Knowledge on the appropriate type of testing to perform may also be limited, especially when professional genetic staffing is unavailable. Furthermore, some phenotypic or extracardiac manifestations may not present until later in childhood, resulting in delayed diagnoses or selection of less informative genetic testing modalities.

Although these limitations for genetic testing exist and access to professional genetic healthcare workers can be limited, echocardiography is a non-invasive and readily available tool at most centers. A child’s first echocardiogram typically establishes the underlying CHD. Most of our model’s predictive performance was already present in the data generated at that first echocardiogram: the images themselves, the measurements, the narrative report and the Fyler codes, together with the child’s sex, age, height and weight to that date. These inputs reached a balanced top-3 accuracy of 0.758 against a chance value of 0.429, suggesting that many genetic syndromes have recognizable cardiac phenotypes at initial evaluation. Accuracy rose further only with inputs that accrue after that visit: the radiology reports added 0.020, an increment whose confidence interval spans zero, and the lifetime ICD-10 list 0.030. Because both accumulate over the clinical course, the accuracy they add comes at the cost of waiting. Models built on routinely captured data may help clinicians direct confirmatory testing, most usefully in centers where genetic testing and genetics professionals are less accessible, and may inform medical management until molecular confirmation is available.

The “undiagnosed” label is inherently provisional because genetic sample and variant re-analyses, and future gene discoveries may eventually yield diagnoses for some children; a model trained on this outcome would therefore learn the current limits of genetic knowledge rather than a stable biological category. The multimodal fusion model was not advantageous compared to LR in predicting common genetic syndromes. However, it was useful in the open-set setting. When a condition was withheld from training entirely, the fusion embedding distinguished affected children from the trained conditions, and placed children who share a causal gene group near one another. Identifying children who fit none of the common syndromes, by contrast, was done better by the logistic regression. Utilization of such models may be particularly valuable for recognizing individuals who would benefit from more extensive genetic evaluation and broader testing, and support gene discovery.

Several limitations exist. The design is retrospective and at a single-center large tertiary referral center, which introduces referral bias, as children with more severe or prenatally diagnosed conditions are more likely to be delivered at, transferred to, or referred to our center. Every patient underwent genetic testing. Therefore, the undiagnosed group comprised children whose testing was non-diagnostic rather than the untested population a referral tool would meet, and the accuracy of the common-syndrome mismatch flag in that untested population is unknown. ICD-10 codes were available only as an undated lifetime list, so performance using them represents an upper bound. ICD-10 codes were also incomplete, available for 91.1% of the cohort and ranging from 44% of children with trisomy 13 to 96% of those with trisomy 21. Children absent from the coded record had fewer echocardiogram reports and fewer Fyler codes overall, so this missingness may be non-random. The Fyler codes are assessments provided by pediatric cardiologists which are subject to inter-provider variability, and genetic diagnoses made before Fyler code assignments may not have been concealed. Five of the seven trained classes are detectable by first-line cytogenetic testing; Kabuki syndrome and the *ELN* sequence variants merged into the Williams class are not. The shortlist’s near-term value is therefore triage and phenocopy recognition, and it does not replace testing. Rare-class sizes are small, and the leave-classes-out intervals rest on 11 classes. A valid growth z-score at the time of the echocardiogram was available only for 61% of the cohort. About 35 confidence intervals are reported without adjustment for multiplicity, as effect estimates for a pre-specified set of comparisons. Of the fourteen effects described as real, five would survive a Bonferroni correction over 35 comparisons: the echocardiogram-report step for both models, the lifetime ICD-10 step for the fusion, the ICD-10 gain over the at-visit model when the notes are omitted, and the coded-only model’s advantage over the full model. The other nine, among them the echocardiographic gain over demographics, the fusion’s imaging gains at the measurements and report stages, the radiology report presence effect, and the +0.051 gain of the full lifetime record over the at-visit model (p = 0.002), would not, and no conclusion rests on them. Finally, echocardiographic results are from one center’s scanners, and transfer of the imaging branch across scanners is untested.

## CONCLUSION

In children with CHD, the information generated by the first echocardiogram together with sex, age, height and weight, carries most of the achievable accuracy for shortlisting the underlying genetic condition with further improvement from the record that accumulates afterwards. A multimodal fusion model did not improve the identification of genetic syndromes over logistic regression, though it recognized rare or unknown conditions that were withheld from training and further grouped children sharing a suspected causal gene group, supporting nearest-neighbor retrieval for a child who remains undiagnosed. Prospective, multi-center evaluation is warranted prior to use of these models for guidance of medical management, counseling and genetic testing.

## Acknowledgments

The EchoPrime software used in this research was created by Milos Vukadinovic, Bryan He, and David Ouyang of Cedars-Sinai Medical Center. © 2024 Cedars-Sinai Medical Center.

## Sources of Funding

This project was supported by OT2 OD038054 (principal investigator: Dr. Aleksandar Milosavljevic) from the National Institutes of Health/Office of the Director.

## Disclosures

None.

## Data Availability

Due to the rarity of CHD diagnoses and the granularity of individual patients’ multimodal input, patient data is not available to request due to the potential of inadvertently re-identifying patients. Code is available on request.

## Notes

### Competing Interest Statement

The authors have declared no competing interest.

### Author Declarations

The Institutional Review Board of Baylor College of Medicine gave ethical approval for this work (protocol H-57914).

## References

1. Pierpont, M. E. et al. Genetic Basis for Congenital Heart Disease: Revisited: A Scientific Statement From the American Heart Association. Circulation 138, e653–e711 (2018).

2. Zaidi, S. & Brueckner, M. Genetics and Genomics of Congenital Heart Disease. Circ. Res. 120, 923–940 (2017).

3. Bigelow, A. M. et al. Genetic Evaluation Practices for Neonates with Congenital Heart Disease in Pediatric Cardiac Intensive Care Units: Findings from a Multi-institutional Survey. Pediatr. Cardiol. https://doi.org/10.1007/s00246-025-04082-2 (2025) doi:10.1007/s00246-025-04082-2.

4. Mustafa, H. J. et al. Yield of Whole Genome Sequencing for Pathogenic Single Nucleotide Variants in Congenital Heart Disease: A Systematic Review and Meta-Analysis. Prenat. Diagn. 46, 780–818 (2026).

5. Durbin, M. D. et al. Rapid Genome Sequencing Shows Diagnostic Utility in Infants With Congenital Heart Defects. Circ. Genomic Precis. Med. 19, e005367 (2026).

6. Keefe, A. C. et al. Implementation of First-Line Rapid Genome Sequencing for Children in Pediatric and Cardiac Intensive Care Units. Am. J. Med. Genet. A. 200, 1326–1336 (2026).

7. Gurovich, Y. et al. Identifying facial phenotypes of genetic disorders using deep learning. Nat. Med. 25, 60–64 (2019).

8. Hsieh, T.-C. et al. GestaltMatcher facilitates rare disease matching using facial phenotype descriptors. Nat. Genet. 54, 349–357 (2022).

9. Dingemans, A. J. M. et al. PhenoScore quantifies phenotypic variation for rare genetic diseases by combining facial analysis with other clinical features using a machine-learning framework. Nat. Genet. 55, 1598–1607 (2023).

10. Murdock, D. R. et al. Early diagnosis of vascular Ehlers-Danlos syndrome through AI-powered facial analysis: Results from the Montalcino Aortic Consortium. Genet. Med. Open 3, 103434 (2025).

11. Porras, A. R., Rosenbaum, K., Tor-Diez, C., Summar, M. & Linguraru, M. G. Development and evaluation of a machine learning-based point-of-care screening tool for genetic syndromes in children: a multinational retrospective study. *Lancet Digit*. Health 3, e635–e643 (2021).

12. Čaplovičová, M. et al. Modeling age-specific facial development in Williams–Beuren-, Noonan-, and 22q11.2 deletion syndromes in cohorts of Czech patients aged 3–18 years: A cross-sectional three-dimensional geometric morphometry analysis of their facial gestalt. Am. J. Med. Genet. A. 176, 2604–2613 (2018).

13. Duong, D. et al. Neural Networks for Classification and Image Generation of Aging in Genetic Syndromes. Front. Genet. **Volume** 13-2022, (2022).

14. Echeverry-Quiceno, L. M. et al. Population-specific facial traits and diagnosis accuracy of genetic and rare diseases in an admixed Colombian population. Sci. Rep. 13, 6869 (2023).

15. Sobreira, N. L. M. et al. Matchmaker Exchange. Curr. Protoc. Hum. Genet. 95, 9.31.1–9.31.15 (2017).

16. Pontikos, N. et al. Next-generation phenotyping of inherited retinal diseases from multimodal imaging with Eye2Gene. *Nat*. Mach. Intell. 7, 967–978 (2025).

17. Vukadinovic, M. et al. Comprehensive echocardiogram evaluation with view primed vision language AI. Nature 650, 970–977 (2026).

18. WHO Child Growth Standards based on length/height, weight and age. Acta Paediatr. Oslo Nor. 1992 Suppl. 450, 76–85 (2006).

19. 2000 *CDC Growth Charts for the United States*. (Public Health Service, Centers for Disease Control and Prevention, National Center for Health Statistics, Hyattsville, Md, 2002).

20. Colan, S. D. Normal echocardiographic values for cardiovascular structures. Echocardiogr. Pediatr. Congenit. Heart Dis. Fetus Adult 883–901 (2016).

21. Sluysmans, T. & Colan, S. D. Structural Measurements and Adjustments for Growth. in Echocardiography in Pediatric and Congenital Heart Disease 61–72 (2016). doi:10.1002/9781118742440.ch5.

22. BCH Z-Score Calculator - Home. https://zscore.chboston.org/.

23. Fyler, D. C. et al. Report of the New England Regional Infant Cardiac Program. Pediatrics 65, 375–461 (1980).

24. Brodersen, K. H., Ong, C. S., Stephan, K. E. & Buhmann, J. M. The Balanced Accuracy and Its Posterior Distribution. in 2010 20th International Conference on Pattern Recognition 3121–3124 (2010). doi:10.1109/ICPR.2010.764.

25. Scheirer, W. J., de Rezende Rocha, A., Sapkota, A. & Boult, T. E. Toward Open Set Recognition. IEEE Trans. Pattern Anal. Mach. Intell. 35, 1757–1772 (2013).

26. McInnes, L., Healy, J. & Melville, J. UMAP: Uniform Manifold Approximation and Projection for Dimension Reduction. Preprint at 10.48550/ARXIV.1802.03426 (2018).

27. Nora, J. J. et al. Risk to offspring of parents with congenital heart defects. JAMA 209, 2052–2053 (1969).

28. Øyen, N. et al. Recurrence of congenital heart defects in families. Circulation 120, 295–301 (2009).

29. Guo, R. et al. Genetic findings of children with congenital heart diseases using chromosomal microarray and trio-based whole exome sequencing. Sci. Rep. 15, 27312 (2025).

30. Meberg, A., Hals, J. & Thaulow, E. Congenital heart defects--chromosomal anomalies, syndromes and extracardiac malformations. Acta Paediatr. Oslo Nor. 1992 96, 1142–1145 (2007).

31. Grech, V. & Gatt, M. Syndromes and malformations associated with congenital heart disease in a population-based study. Int. J. Cardiol. 68, 151–156 (1999).

32. Ferencz, C., Boughman, J. A., Neill, C. A., Brenner, J. I. & Perry, L. W. Congenital cardiovascular malformations: questions on inheritance. Baltimore-Washington Infant Study Group. J. Am. Coll. Cardiol. 14, 756–763 (1989).

33. Rodriguez, K. M. et al. Rapid Whole-Genome Sequencing and Clinical Management in the PICU: A Multicenter Cohort, 2016-2023. Pediatr. Crit. Care Med. J. Soc. Crit. Care Med. World Fed. Pediatr. Intensive Crit. Care Soc. 25, 699–709 (2024).

34. Kikano, S. et al. Postoperative Morbidity and Interstage Hemodynamics Following Stage I Palliation in Patients with Turner Syndrome and Hypoplastic Left Heart Syndrome. Pediatr. Cardiol. 45, 221–227 (2024).

35. Nugent, D. J., Romano, A. A., Sabharwal, S. & Cooper, D. L. Evaluation of bleeding disorders in patients with Noonan syndrome: a systematic review. J. Blood Med. 9, 185–192 (2018).

36. Collins, R. T. 2nd. Cardiovascular disease in Williams syndrome. Circulation 127, 2125–2134 (2013).

37. Bird, L. M. et al. Sudden death in Williams syndrome: report of ten cases. J. Pediatr. 129, 926–931 (1996).

38. Horowitz, P. E., Akhtar, S., Wulff, J. A., Al Fadley, F. & Al Halees, Z. Coronary artery disease and anesthesia-related death in children with Williams syndrome. J. Cardiothorac. Vasc. Anesth. 16, 739–741 (2002).

39. Wessel, A. et al. Risk of sudden death in the Williams-Beuren syndrome. Am. J. Med. Genet. A. 127A, 234–237 (2004).

40. Marino, B. S. et al. Neurodevelopmental outcomes in children with congenital heart disease: evaluation and management: a scientific statement from the American Heart Association. Circulation 126, 1143–1172 (2012).

41. Gaynor, J. W. et al. Patient characteristics are important determinants of neurodevelopmental outcome at one year of age after neonatal and infant cardiac surgery. J. Thorac. Cardiovasc. Surg. 133, 1344–1353, 1353.e1–3 (2007).

42. Kaltman, J. R. et al. Neurodevelopmental outcome after early repair of a ventricular septal defect with or without aortic arch obstruction. J. Thorac. Cardiovasc. Surg. 131, 792–798 (2006).

43. Zeltser, I. et al. Genetic factors are important determinants of neurodevelopmental outcome after repair of tetralogy of Fallot. J. Thorac. Cardiovasc. Surg. 135, 91–97 (2008).

44. Atallah, J. et al. Two-year general and neurodevelopmental outcome after neonatal complex cardiac surgery in patients with deletion 22q11.2: a comparative study. J. Thorac. Cardiovasc. Surg. 134, 772–779 (2007).

