## Supplemental Tables 1-3 for "Artificial Intelligence-Based Modeling to Aid in the Identification of Genetic Conditions in Infants with Congenital Heart Disease"

### Supplemental Table 1. Echocardiographic Measurements Obtained From Echocardiogram Reports

**Supplemental Table 1.** Measurements obtained from echocardiogram reports.

| Variable | Unit | Echocardiogram View |
|---|---|---|
| Aortic valve annulus* | cm | PLAX |
| Aortic root* | cm | PLAX |
| Aortic sinotubular junction* | cm | PLAX |
| Ascending aorta* | cm | PLAX |
| Proximal transverse arch | cm | SSN |
| Distal transverse arch* | cm | SSN |
| Aortic isthmus* | cm | SSN |
| Pulmonary valve annulus* | cm | PSAX |
| Main pulmonary artery* | cm | PSAX |
| Left pulmonary artery* | cm | PSAX |
| Right pulmonary artery* | cm | PSAX |
| Mitral valve annulus* | cm | A4C |
| Tricuspid valve annulus* | cm | A4C |
| Mitral valve E wave* | m/sec | A4C |
| Mitral valve A wave* | m/sec | A4C |
| IVS end-diastolic, MM* | cm | PSAX, MM |
| IVS end-systolic, MM* | cm | PSAX, MM |
| LV PW end-diastolic, MM* | cm | PSAX, MM |
| LV PW end-systolic, MM* | cm | PSAX, MM |
| LV end-diastolic diameter, MM* | cm | PSAX, MM |
| LV end-systolic diameter, MM* | cm | PSAX, MM |
| LV diastolic dimension, long axis* | cm | A4C |
| LV systolic dimension, long axis* | cm | A4C |
| LV mass, MM* | g | PSAX, MM |
| LV mass indexed to BSA, MM | $g/m^2$ | PSAX, MM |
| LV mass to height | $g/m^{2.7}$ | PSAX, MM |
| Systolic BP | mmHg | N/A |
| Diastolic BP | mmHg | N/A |
| Mean BP | mmHg | N/A |
| Pulse pressure | mmHg | N/A |

*Z-score derived from Boston Children's nomograms (Boston Children's Hospital, 2015)
A4C: apical four chamber; BP: blood pressure; BSA: body surface area; cm: centimeters; g: grams; IVS: interventricular septum; LV: left ventricle; m: meters; MM: M-mode; N/A: not applicable; PLAX: parasternal long axis; PSAX: parasternal short axis; PW: posterior wall; SSN: suprasternal notch.

**Supplemental Table 2. Gene-level membership of the merged rare-diagnosis groups used in the leave-one-group-out analysis**

| Group | n | Original diagnosis labels (n) |
|---|---|---|
| RASopathy | 15 | PTPN11 (7); RAF1 (2); NF1 (2); RIT1 (1); KRAS (1); SOS1 (1); BRAF (1) |
| Ciliary/laterality | 17 | NODAL (6); MMP21 comp het (3); DNAH5/GDF1 digenic (1); DNAH11 comp het (1); PKHD1 comp het (1); CCDC103 homo (1); DNAAF5 homo (1); MKKS comp het (1); DNAH5 comp het (1); GDF1 comp het/AD (1) |
| CHARGE-CHD7 | 17 | CHARGE-CHD7 (17) |
| Alagille | 7 | Alagille (7) |

RASopathy pools any rare diagnosis naming a RAS-pathway gene (PTPN11, SOS1, SOS2, RAF1, BRAF, MAP2K1, MAP2K2, KRAS, NRAS, HRAS, RIT1, RRAS, RRAS2, MRAS, NF1, CBL, SHOC2, A2ML1, SPRED1); ciliary/laterality pools CCDC103, DNAAF5, DNAH11, DNAH5, NODAL, GDF1, MKKS, MMP21 and PKHD1. Groups with seven or more children joined the 11-class panel. Of the 231 rare children, 121 fall in 28 confirmed groups of two or more members and are eligible for the same-cause-neighbor measure; 110 are singletons.

**Supplemental Table 3. Genetic Diagnosis Labels**

| Group | Genetic Diagnosis Label | n |
|---|---|---|
| Common-7 | Trisomy 21 | 580 |
| Common-7 | 22q11.2 deletion syndrome | 152 |
| Common-7 | Williams syndrome and *ELN* variants | 91 |
| Common-7 | Trisomy 18 | 67 |
| Common-7 | Turner syndrome, monosomy X | 45 |
| Common-7 | Trisomy 13 | 34 |
| Common-7 | Kabuki syndrome-*KMT2D* | 19 |
| Rare diagnosis | CHARGE syndrome-*CHD7* | 17 |
| Rare diagnosis | *PTPN11*(RASopathy group) | 7 |
| Rare diagnosis | Alagille syndrome (*JAG1 and NOTCH 2*) | 7 |
| Rare diagnosis | *NODAL* | 6 |
| Rare diagnosis | *NOTCH1* | 6 |
| Rare diagnosis | 1p36 deletion syndrome | 5 |
| Rare diagnosis | 1q21.1 deletion | 5 |
| Rare diagnosis | Turner syndrome, mosaic | 4 |
| Rare diagnosis | *FLT4* | 4 |
| Rare diagnosis | 8p23.1 deletion, involving *GATA4* | 3 |
| Rare diagnosis | *ZEB2* | 3 |
| Rare diagnosis | 1q21.1q21.2 duplication | 3 |
| Rare diagnosis | *MMP21* compound heterozygote | 3 |
| Rare diagnosis | *MYRF* | 3 |
| Rare diagnosis | *SMAD2* | 3 |
| Rare diagnosis | *MED13L* syndrome | 2 |
| Rare diagnosis | 1q21.1 duplication | 2 |
| Rare diagnosis | 3q29 deletion | 2 |
| Rare diagnosis | Trisomy 22 | 2 |
| Rare diagnosis | *RAF1* | 2 |
| Rare diagnosis | *GATA4* | 2 |
| Rare diagnosis | Trisomy 22, Mosaic | 2 |
| Rare diagnosis | Male gonadal dysgenesis (mosaic XY, XO) | 2 |
| Rare diagnosis | *NF1* (RASopathy group) | 2 |
| Rare diagnosis | 22q11 duplication | 2 |
| Rare diagnosis | 16p11.2 duplication syndrome | 2 |
| Rare diagnosis | *NKX2.5* | 2 |
| Rare diagnosis | TEK exonic deletion | 2 |
| Rare diagnosis | 2q37 deletion | 2 |
| Rare diagnosis | *CTNND1* | 2 |
| Rare diagnosis | *PKP2* | 1 |
| Rare diagnosis | 9q34.3 del (Kleefstra syndrome) and 5q35.2q35.3 duplication | 1 |

| | | |
|---|---|---|
| Rare diagnosis | 4q35.1 deletion | 1 |
| Rare diagnosis | *RIT1* (RASopathy group) | 1 |
| Rare diagnosis | 15q11.2 deletion, *FBLN2* | 1 |
| Rare diagnosis | Kabuki-*KMT2D*, 15q11.2 duplication | 1 |
| Rare diagnosis | *DNAH5/GDF1* digenic (ciliary group) | 1 |
| Rare diagnosis | 16p13.3 duplication | 1 |
| Rare diagnosis | 16p13.11 duplication | 1 |
| Rare diagnosis | 8p23.1 duplication (including *GATA4*) AND 8p23.2p23.1 duplication | 1 |
| Rare diagnosis | Trisomy 13, Robertsonian translocation | 1 |
| Rare diagnosis | *TTC21B* compound heterozygote | 1 |
| Rare diagnosis | 3p26.3p26.2 del, 16p13.3p13.2 dup (unbalanced translocation) | 1 |
| Rare diagnosis | *CNOT9* | 1 |
| Rare diagnosis | 20q13.13q13.2 duplication | 1 |
| Rare diagnosis | Myhre syndrome-*SMAD4* | 1 |
| Rare diagnosis | *TPM1* | 1 |
| Rare diagnosis | *KRAS* (RASopathy group) | 1 |
| Rare diagnosis | 22q11.21 duplication + *EVC2* homozygous | 1 |
| Rare diagnosis | 8q12.1q12.2 deletion, involving CHD7 | 2 |
| Rare diagnosis | *TTC25* | 1 |
| Rare diagnosis | 15q26-qter deletion syndrome | 1 |
| Rare diagnosis | 1q23.3q32.1 deletion | 1 |
| Rare diagnosis | *TOP2B* | 1 |
| Rare diagnosis | *NOMO3* | 1 |
| Rare diagnosis | Opitz GBB syndrome | 1 |
| Rare diagnosis | 17p12 deletion | 1 |
| Rare diagnosis | Kabuki-*KDM6A* | 1 |
| Rare diagnosis | *DNAH11* compound heterozygote (ciliary group) | 1 |
| Rare diagnosis | Xp22.21p11.3 deletion | 1 |
| Rare diagnosis | 13q12.2q14.2 deletion | 1 |
| Rare diagnosis | PKHD1 compound heterozygote (ciliary group) | 1 |
| Rare diagnosis | *TNNT2* | 1 |
| Rare diagnosis | Trisomy 14 | 1 |
| Rare diagnosis | 22q11.2 distal duplication | 1 |
| Rare diagnosis | *NAA15* | 1 |
| Rare diagnosis | Kleefstra type II- *KMT2C* | 1 |
| Rare diagnosis | *SMAD6* | 1 |
| Rare diagnosis | Williams syndrome AND *KRIT1* | 1 |
| Rare diagnosis | Cornelia De Lange Syndrome-*NIPBL* | 1 |
| Rare diagnosis | *EFTUD2* | 1 |
| Rare diagnosis | *ARHGAP35* | 1 |
| Rare diagnosis | 1q42.12 duplication | 1 |

| | | |
|---|---|---|
| Rare diagnosis | NADSYN1 homo | 1 |
| Rare diagnosis | 17p11.2 deletion-Smith Magenis syndrome | 1 |
| Rare diagnosis | Cornelia De Lange Syndrome -CDLS2-Xlinked | 1 |
| Rare diagnosis | 8p23.1 deletion, involving GATA4; also duplication within the pseudoautosomal (PAR1) region of chromosomes X (Xp22.33) and Y (Yp11.32) involving the SHOX gene | 1 |
| Rare diagnosis | *FOXF1* | 1 |
| Rare diagnosis | *NR2F2* | 1 |
| Rare diagnosis | 8p23.1 duplication including *GATA4* | 1 |
| Rare diagnosis | 15q11.2 deletion | 2 |
| Rare diagnosis | Tatton-Brown-Rahman Syndrome-*DNMT3A* | 1 |
| Rare diagnosis | 17q21.31 deletion | 1 |
| Rare diagnosis | CCDC103 homozygote (ciliary group) | 1 |
| Rare diagnosis | 16p11.2 deletion | 1 |
| Rare diagnosis | 16p13.11 deletion | 1 |
| Rare diagnosis | Turner syndrome and 1q21.1q21.2 dup | 1 |
| Rare diagnosis | 18p11.2 deletion | 1 |
| Rare diagnosis | 22q11.2 deletion syndrome with unbalanced Translocation between 12 and 22 | 1 |
| Rare diagnosis | *SOS1*(RASoapthy group) | 1 |
| Rare diagnosis | 18q deletion syndrome | 1 |
| Rare diagnosis | *ACVR2B* | 1 |
| Rare diagnosis | *FGFR1* | 1 |
| Rare diagnosis | *MYBC3* | 1 |
| Rare diagnosis | 4q25335.2 duplication (partial trisomy 4q), 20p13 del | 1 |
| Rare diagnosis | SMARCA4 | 1 |
| Rare diagnosis | ACVRL1 | 1 |
| Rare diagnosis | Unbalanced 10;21 translocation causing partial trisomy of 10p and 10q and partial monosomy of 21q | 1 |
| Rare diagnosis | 22q11.21 distal deletion | 1 |
| Rare diagnosis | 8p tetrasomy (mosaic) | 1 |
| Rare diagnosis | Trisomy 21 Mosaic | 1 |
| Rare diagnosis | *NKX2-6* | 1 |
| Rare diagnosis | *DNAAF5* homo | 1 |
| Rare diagnosis | Trisomy 8 mosaic | 1 |
| Rare diagnosis | *TAB2* | 1 |
| Rare diagnosis | unbalanced translocation between chromosomes 9q and 17q (trisomy of distal 17q) | 1 |
| Rare diagnosis | 7q31.3 deletion | 1 |

| | | |
|---|---|---|
| Rare diagnosis | *FBLN5* (homozygous), autosomal recessive cutis laxa type 1b (ARCL1b) | 1 |
| Rare diagnosis | Trisomy 18 Mosaic | 1 |
| Rare diagnosis | *MKKS* compound heterozygote (ciliary group) | 1 |
| Rare diagnosis | Xp21.1 duplication, including *DMD* gene | 1 |
| Rare diagnosis | 6q15q21 deletion | 1 |
| Rare diagnosis | Pallister-Killian syndrome -Tetrasomy 12p mosaic | 1 |
| Rare diagnosis | 3pter-p25 deletion syndrome | 1 |
| Rare diagnosis | *MYH7* | 1 |
| Rare diagnosis | *TBX2* | 1 |
| Rare diagnosis | *ACADM* compound heterozygote | 1 |
| Rare diagnosis | 1q32.2-1q44 duplication (partial trisomy) and monosomic for the terminal end of 1q44 | 1 |
| Rare diagnosis | 4p16.3p15.33 duplication | 1 |
| Rare diagnosis | 4p16.3q22.2 duplication including the entire short (p) arm, the centromere, and the proximal long (q) arm of chromosome 4. In addition, 18q23 duplication, suggestive an unbalanced translocation | 1 |
| Rare diagnosis | 11q24.2q25 deletion, Jacobsen syndrome | 1 |
| Rare diagnosis | XYY | 1 |
| Rare diagnosis | 1q44 distal deletion | 1 |
| Rare diagnosis | 18p tetrasomy mosaic and a small part of 18q11.2 ; 18p-mosaicism for a copy number gain of 18p and proximal 18q through band 18q11.2 | 1 |
| Rare diagnosis | 7q21.3q31.1 deletion | 1 |
| Rare diagnosis | 15q26.2q26.3 deletion | 1 |
| Rare diagnosis | *DNAH5* compound heterozygote (ciliary group) | 1 |
| Rare diagnosis | *LEFTY* | 1 |
| Rare diagnosis | *BRAF (RASopathy group)* | 1 |
| Rare diagnosis | 8p23.3p23.1 del AND 8p23.1 dup (including GATA4) AND 8p23.1p21.3 dup | 1 |
| Rare diagnosis | *EPHB4* | 1 |
| Rare diagnosis | 4q27q35.2 duplication, 4q35.2 deletion | 1 |
| Rare diagnosis | 15q13.3 duplication | 1 |
| Rare diagnosis | T13 Mosaic | 1 |
| Rare diagnosis | GDF1 compound heterozygote | 1 |
| Rare diagnosis | Trisomy 16 and 11q deletion | 1 |
| Rare diagnosis | Chr 6 ring including deletion of both the short (p) and long (q) arms, as well as a mosaic duplication of remaining 6p25.3q26 region | 1 |

| | | |
|---|---|---|
| Rare diagnosis | 16p11.2q24.3 duplication including the entire long (q) arm, the centromere and the pericentromeric region of short arm | 1 |
| Rare diagnosis | 5p15.32 del | 1 |
| Rare diagnosis | 1p36.32 to 1p36.33 dup | 1 |
| Rare diagnosis | ATP5F1A; 2 mitochondrial PVs, 1 os AD | 1 |
| Rare diagnosis | *IFT122* | 1 |
| Rare diagnosis | *ANKRD11* | 1 |
| Rare diagnosis | *LZTR1* | 1 |
| Rare diagnosis | 10q22.3q23.3 deletion | 1 |
| Rare diagnosis | 15q11.2 duplication | 1 |
| Rare diagnosis | 8q22.1q24.3 dup AND 11q25 del (unbalanced translocation) | 1 |
| Rare diagnosis | TBX5 | 1 |
| Rare diagnosis | 9p partial trisomy and partial monosomy of 18q; 46,XY,der(18)t(9;18)(p21.3;q22.3) - Unbalanced translocation | 1 |
| Rare diagnosis | 14q24.3q32.33:Uniparenteral disomy-Temple Syndrome | 1 |
| Undiagnosed | Unsolved | 501 |
